# The NASSS (Non-Adoption, Abandonment, Scale-Up, Spread and Sustainability) framework use over time: A scoping review

**DOI:** 10.1101/2023.11.22.23298897

**Authors:** Hwayeon Danielle Shin, Emily Hamovitch, Evgenia Gatov, Madison MacKinnon, Luma Samawi, Rhonda Boateng, Kevin Thorpe, Melanie Barwick

**Affiliations:** Institute of Health Policy, Management, and Evaluation, University of Toronto, Toronto, Canada; Krembil Centre for Neuroinformatics, Centre for Addiction and Mental Health, Toronto, Canada; The Centre for Addiction and Mental Health, Toronto, Canada; Child Health Evaluative Sciences, The Peter Gilgan Centre for Research and Learning, The Hospital for Sick Children, Toronto, ON, Canada; Department of Psychiatry, University of Toronto, Toronto, ON, Canada

**Author notes:** ***Correspondence to:*** Hwayeon Danielle Shin, RN MScN PhD(c).

**Keywords:** Scoping Review, Implementation Science, NASSS

## Abstract

**Background:** The Non-adoption, Abandonment, Scale-up, Spread, Sustainability (NASSS) framework (2017) was established as an evidence-based, theory-informed tool to predict and evaluate the success of implementing health and care technologies. While the NASSS is gaining popularity, its use has not been systematically described. Literature reviews on the applications of popular implementation frameworks such as RE-AIM and CFIR have enabled their advancement in the implementation science field. Similarly, we sought to advance the science of implementation and application of theories, models, and frameworks (TMFs) in research by exploring the application of the NASSS in the five years since its inception.

**Objective:** We aim to understand the characteristics of studies that used the NASSS, how it was used, and the lessons learned from its application.

**Methods:** We conducted a scoping review following the Joanna Briggs Institute methodology. We searched the following databases on December 20, 2022: Ovid MEDLINE, EMBASE, PsychINFO, CINAHL, Scopus, Web of Science, and LISTA. We used typologies and frameworks to characterize evidence to address our aim.

**Results:** This review included 57 studies, which were a mix of qualitative (n=28), mixed/multi-methods (n=13), case studies (n=6), observational (n=3), experimental (n=3), and other designs (e.g., quality improvement) (n=4). The four most common types of digital applications being implemented were telemedicine/virtual care (n=24), personal health devices (n=10), digital interventions, such as internet Cognitive Behavioural Therapies (n=10), and knowledge generation applications (n=9). Studies used the NASSS to inform study design (n=9), data collection (n=35), analysis (n=41), data presentation (n=33), and interpretation (n=39). Most studies applied the NASSS retrospectively to implementation (n=33). The remainder applied the NASSS prospectively (n=15) or concurrently (n=8) with implementation. We also collated reported barriers and enablers to implementation. We found the most reported barriers fell within the Organization and Adopter System domains, and the most frequently reported enablers fell within the Value Proposition domain. Eighteen studies highlighted the NASSS as a valuable and practical resource, particularly for unravelling complexities, comprehending implementation context, understanding contextual relevance in implementing health technology, and recognizing the NASSS’ adaptable nature to cater to researchers’ requirements.

**Conclusions:** Most studies used the NASSS retrospectively, which may be attributed to the framework’s novelty. However, this finding highlights the need for prospective and concurrent application of the NASSS within the implementation process. In addition, almost all included studies reported multiple domains as barriers and enablers to implementation, indicating that implementation is a highly complex process that requires careful preparation to ensure implementation success. Finally, we identified a need for better reporting when using the NASSS in implementation research to contribute to the collective knowledge in the field.

## Introduction

Healthcare technology innovations hold considerable promise for enhancing patient outcomes and service efficiency, but they frequently remain confined to small-scale demonstration initiatives [1–5]. Moreover, current evidence indicates a prevalent pattern of non-adoption and abandonment of healthcare technology innovations by their intended users, with limited success in integrating these innovations into regular practice or expanding their implementation to different contexts [6]. This challenge is especially evident in complex healthcare settings, where the multifaceted nature of the innovations and the environment can create barriers to successful implementation [7].

Healthcare is described as a complex adaptive system, discouraging simplistic linear cause-and-effect reasoning [8,9]. Instead, there is a growing recognition of the need to emphasize dynamic processes while implementing healthcare practices. This change in perspective reflects an understanding that healthcare is influenced by multifaceted interactions and feedback loops that cannot be adequately explained by linear models alone. In response to this evolving perspective, the Non-Adoption, Abandonment, Scale-up, Spread, and Sustainability (NASSS) framework was introduced in 2017 [10]. NASSS was developed as an evidence-based and theory-informed approach to enhance the ability to predict and assess the success of implementing innovative technologies in the healthcare context [10]. Related complexity assessment tools (NASSS-CAT) were developed in 2020 to enhance understanding, guide monitoring, and facilitate research on technology projects in healthcare or social care settings through stakeholder discussions [11].

The NASSS encompasses seven distinct domains: 1) Illness/Condition; 2) Technology; 3) Value Proposition; 4) Adopter System; 5) Organization(s); 6) Wider Context; and 7) Embedding and Adaptation Over Time [10]. Each domain can be categorized as simple, complicated, or complex [10]. The greater the complexity observed within these domains, the more obstacles will likely arise, hindering the successful adoption, scale-up, spread, and sustainability of innovative health and care technologies [10]. The NASSS framework considers the intricate web of dynamic interactions that influence the adoption and outcomes of innovations and aims to provide a more comprehensive and accessible tool for evaluating and improving the implementation of healthcare innovations [10].

Although new, the NASSS framework has been well-received. The seminal paper has had nearly 750 citations at the time of writing, as reported in the *Journal of Medical Internet Research* [10]. The surge in interest reflects the widespread adoption of the NASSS, which has been utilized prospectively and retrospectively to assess patient-oriented technologies and tools for decision-making purposes [12,13]. Despite its popularity, there has been a lack of systematic documentation regarding the use of the NASSS framework following its release. Likewise, a comprehensive analysis of the framework’s contributions and the insights derived from its application has not been conducted systematically.

The applications of popular implementation theories, models, and frameworks (TMFs), such as the Reach, Effectiveness, Adoption, Implementation, and Maintenance (RE-AIM) and Consolidated Framework for Implementation Research (CFIR), have been well documented in the literature. For example, there have been several literature reviews [14,15] on using RE-AIM since its inception in 1999.

These reviews have described and assessed the application of the RE-AIM and have enabled the advancement of the framework (i.e., enhanced RE-AIM/Pragmatic Robust Implementation and Sustainability Model (PRISM) 2019) as well as its novel application, such as an opportunity to use the RE-AIM in combination with the Pragmatic Explanatory Continuum Indicator Summary (PRECIS) model [14,15].

Similarly, we aim to contribute to the field of implementation science by exploring the NASSS applications to date and identifying opportunities to advance the framework. A scoping review is the selected method and was deemed most appropriate because our primary objective is to provide a breadth of literature currently available on the NASSS application [16]. A preliminary search of PROSPERO, MEDLINE, the Cochrane Database of Systematic Reviews, Open Science Framework, and *JBI Evidence Synthesis* was conducted in October 2022. No current or in-progress scoping or systematic reviews on the topic were identified.

## Review questions

1. What are the characteristics of studies that used the NASSS?
2. How has the NASSS been used in the identified studies, including, but not limited to, timing within implementation, depth of application, and use in combination with other tools (e.g., the NASSS-CAT)?
3. What are the author-reported lessons learned from applying the NASSS?

## Inclusion criteria

### Concept

This review included all studies that used the NASSS framework and/or NASSS-CAT in their design. Studies that only referred to the framework without application (e.g., citing in the introduction and/or discussion) were excluded.

### Context and population

There were no exclusion criteria for population and context. Any studies conducted in any context with any population were considered for inclusion. However, due to the available resources in our research team, only English-language publications were included.

### Type of sources

This review included all research designs (e.g., quantitative, observational, qualitative, and mixed methods). We also considered peer-reviewed and grey literature, including conference proceedings and dissertations, but we included only empirical studies. Reference lists in non-empirical literature (e.g., reviews) were screened to identify relevant primary studies. Only literature published since 2017, the year of the publication of the seminal NASSS framework paper, was included.

## Methods

This scoping review was conducted following the Joanna Briggs Institute (JBI) methodology for scoping reviews [17,18], and the manuscript was prepared in line with the Preferred Reporting Items for Systematic Reviews and Meta-Analyses extension for Scoping Reviews (PRISMA-ScR) [19]. Our *a priori* protocol [20] was registered on the Open Science Framework.

### Search strategy

In collaboration with a health sciences librarian and following the Peer Review of Electronic Search Strategies (PRESS) guideline [21], a comprehensive search strategy was developed to locate relevant scholarly literature using multiple bibliographic databases. This scoping review followed a three-step search strategy outlined in the JBI methodology. Firstly, an initial limited search of MEDLINE was undertaken to identify articles on the topic. Secondly, the text words in the titles and abstracts of relevant articles and the index terms used to describe the articles were used to develop a complete search strategy. Then, the entire search strategy, including all identified keywords and index terms, was adapted for each included information source and our search was undertaken on December 20, 2022, on the following databases: Ovid MEDLINE, EMBASE, PsychINFO, CINAHL, Scopus, Web of Science, and Library, Information Science and Technology Abstracts (LISTA). Thirdly, reference lists of relevant reviews were screened to identify eligible empirical studies. The full search strategies are provided in Supporting Information 1. Since the NASSS framework was first published in 2017, databases have been searched from 2017 onwards. In addition to a scholarly database search, a forward citation search [22] was used in Scopus and Web of Science on October 13 and 17, 2022, to complement our database searches. The main steps in this forward citation search included using citation indexes to identify studies that cite the original NASSS paper published in 2017. This search strategy helped to identify papers that our search strategy might have missed.

### Study/Source of evidence selection

Following the search, all identified records were collated and uploaded into the Covidence [23], and duplicates were automatically removed. Then, five random articles were selected for our pilot testing, and all five reviewers on the team independently assessed the titles and abstracts against the inclusion criteria. While our pilot testing generally went smoothly, we encountered a need for clarification regarding what constitutes NASSS application. After a team discussion, we clarified that simply citing the NASSS was insufficient for inclusion; instead, the work should incorporate the NASSS or NASSS-CAT tool into some aspect of the study design to ensure consistency in our screening decision-making process, which is often not mentioned in the abstract. Therefore, the team decided to err on the side of caution during the screening phase. After pilot testing for the calibration exercise, the remaining titles and abstracts were screened by sets of two independent reviewers (HDS, EG, MM, EH, LS, RB). Potentially relevant papers were retrieved in full, and their citation details were imported into the Covidence [23]. Two independent reviewers assessed full texts (HDS, EG, MM, EH, LS, RB). Full-text studies that did not meet the inclusion criteria were excluded, and reasons for their exclusion were documented. Any reviewer disagreements were resolved through discussion or with a third reviewer. Scoping reviews typically do not necessitate methodological evaluation [18]; therefore, critical appraisal was omitted.

### Data extraction

Data were extracted from papers by sets of two independent reviewers (HDS, EH, EG, MM, RB, LS) using a data extraction tool developed in collaboration with the research team. We extracted the following information: general characteristics of the paper, intervention characteristics, description of the NASSS framework application, reported implementation barriers and facilitators, and study conclusion and author-reported lessons learned from applying the NASSS. Any reviewer disagreements were resolved through discussion or with a third reviewer. See Supporting Information 2 for our data extraction tool.

### Data analysis and presentation

A descriptive, analytical approach was used to generate summary statistics (e.g., frequency counts, percentages, etc.) for the data extracted concerning the general characteristics of the included studies. Subsequently, a content analysis was conducted to characterize the narrative data. First, the digital applications implemented in the included studies were categorized by two reviewers (MM, HDS) by adapting the framework, ’Evolving Applications of Digital Technology in Health and Health Care.’ Application categories [24] are as follows: 1) Telemedicine/Virtual care; 2) Personal health devices; 3) Digital interventions; 4) Knowledge generation and/or integrators; 5) Health information; 6) Surgical/Radio graphic interventions; 7) Diagnostic and imaging [24]. One innovation could be characterized by more than one category. Secondly, two reviewers categorized health conditions being examined in the included studies into disease types (EH, HDS). Thirdly, the description of the NASSS application was assessed by sets of two independent reviewers (HDS, EH, EG, MM, RB, LS) in terms of its timing within the implementation (i.e., prospective, retrospective, concurrent) and study design aspects (e.g., overall design, data collection, data analysis). This process required some level of interpretation by the team, and any conflicts in interpretation were resolved through discussion. Fourth, barriers and enablers, often correspondingly reported to the primary NASSS domains, were collated from the papers. Then, sets of two reviewers (HDS, EH, EG, MM, RB) categorized these into subdomains of NASSS. Fifth, reported lessons learned from the authors were narratively summarized. The charted results are accompanied by narrative summaries that describe how the results relate to our review objectives and questions.

## Results

Our search strategy yielded 1,705 citations (Figure 1). Following the automatic removal of duplicates by Covidence, 823 articles underwent title and abstract screening, and 355 articles underwent full-text evaluation to culminate in 57 studies in this review. Most excluded studies cited the NASSS framework in the text (e.g., in the discussion) but did not use the framework in study design, data collection, analyses, or presentation of results. Other excluded studies were non-empirical (e.g., commentary) and those for which full text was unavailable.

**Figure 1:**
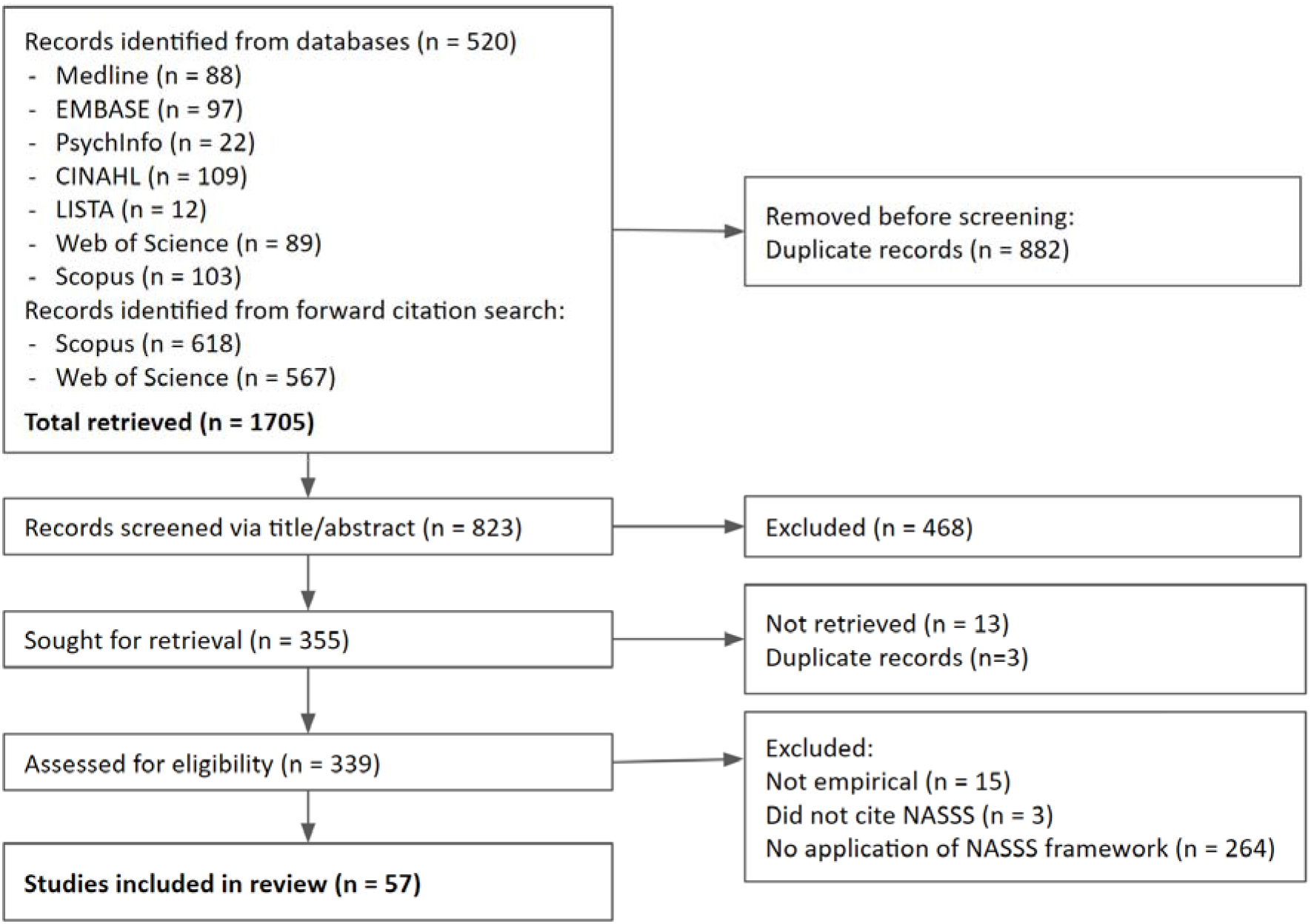
PRISMA Flow Diagram

### RQ1. Characteristics of included studies

Individual study characteristics are presented in Table 1. As indicated in summary Table 2, among the 57 included studies, the majority were qualitative (n=28), following mixed/multi-methods (n=13), case-studies (n=6), observational (n=3), experimental (n=3), and other designs (e.g., quality improvement, n=4). Many studies originated in the United Kingdom (n=15), Australia (n=13), and the United States (n=9), with a few other studies being set elsewhere in Europe, Southeast Asia, and North America. However, It is noteworthy that the NASSS framework was developed in the United Kingdom, and several included studies were part of the initial empirical testing and refinement of the NASSS domains [25].

**Table 1.**
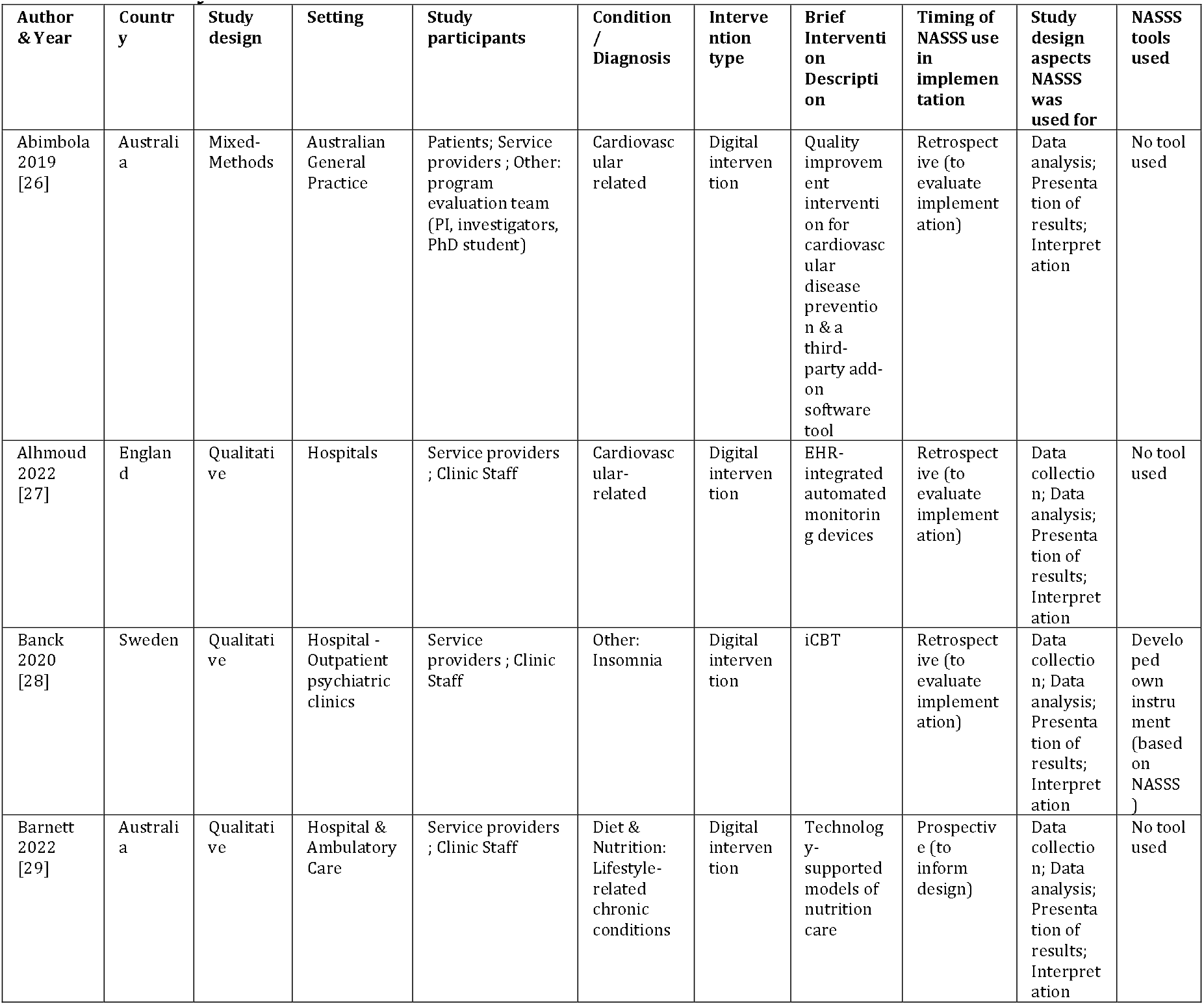
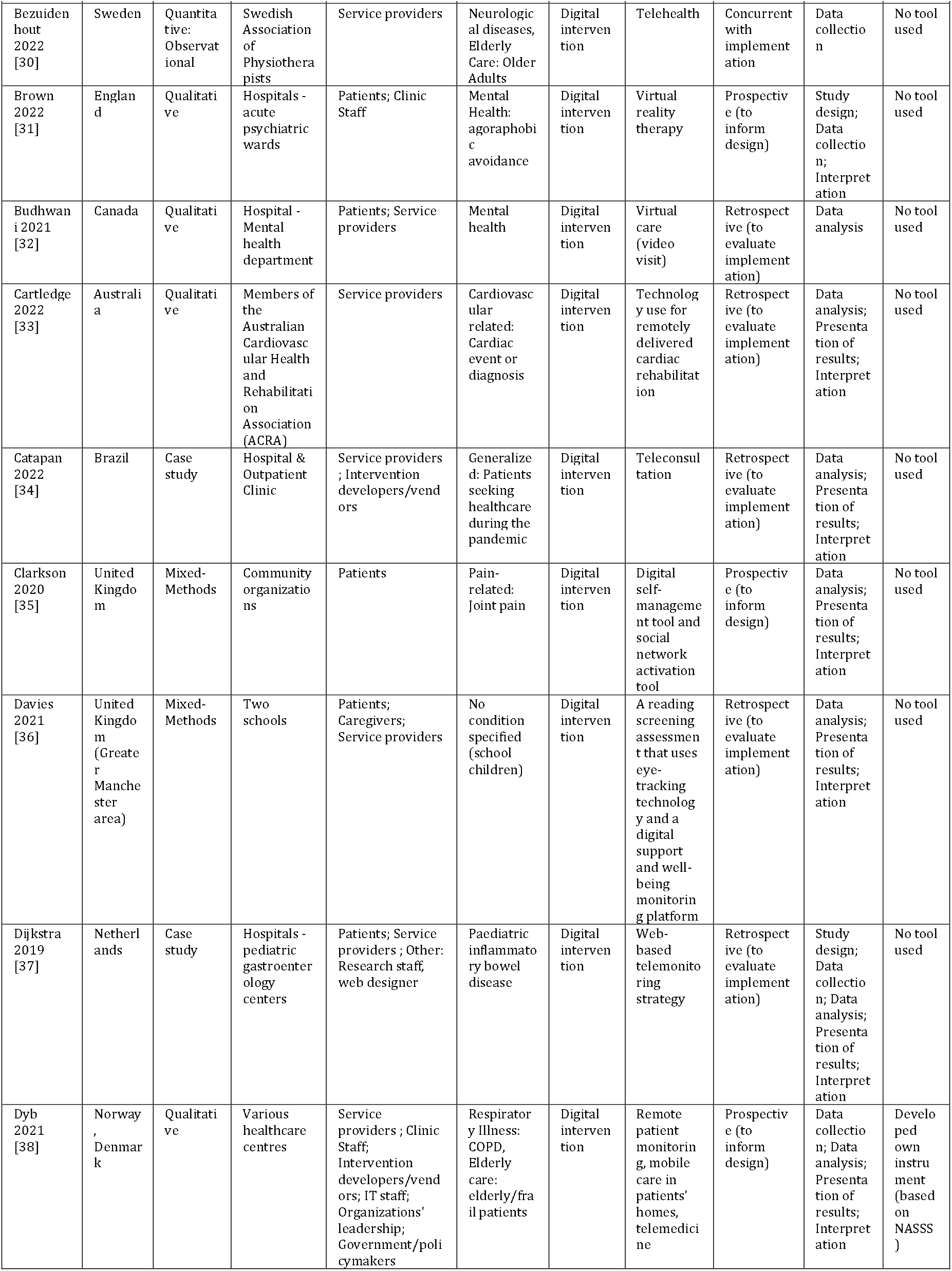
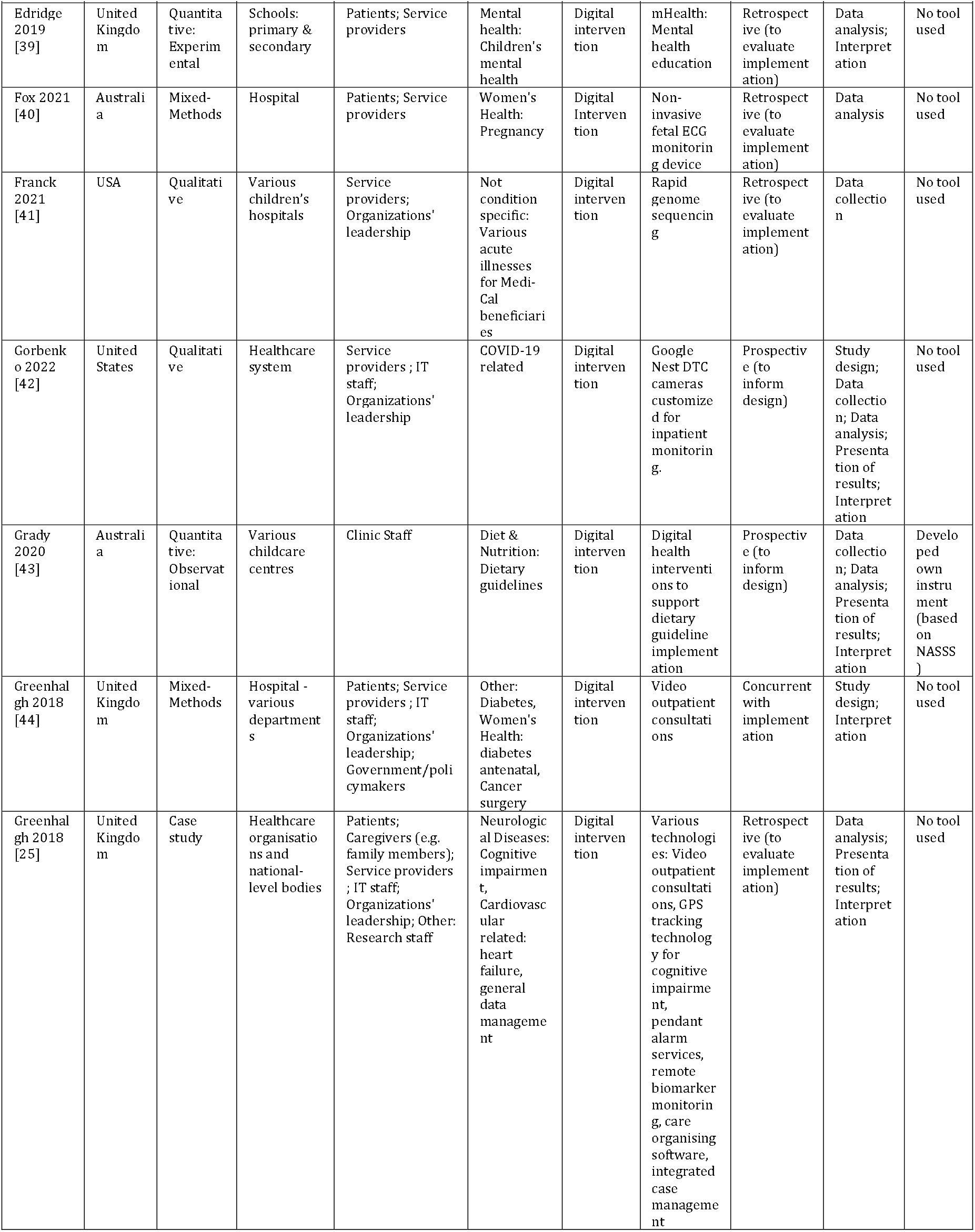
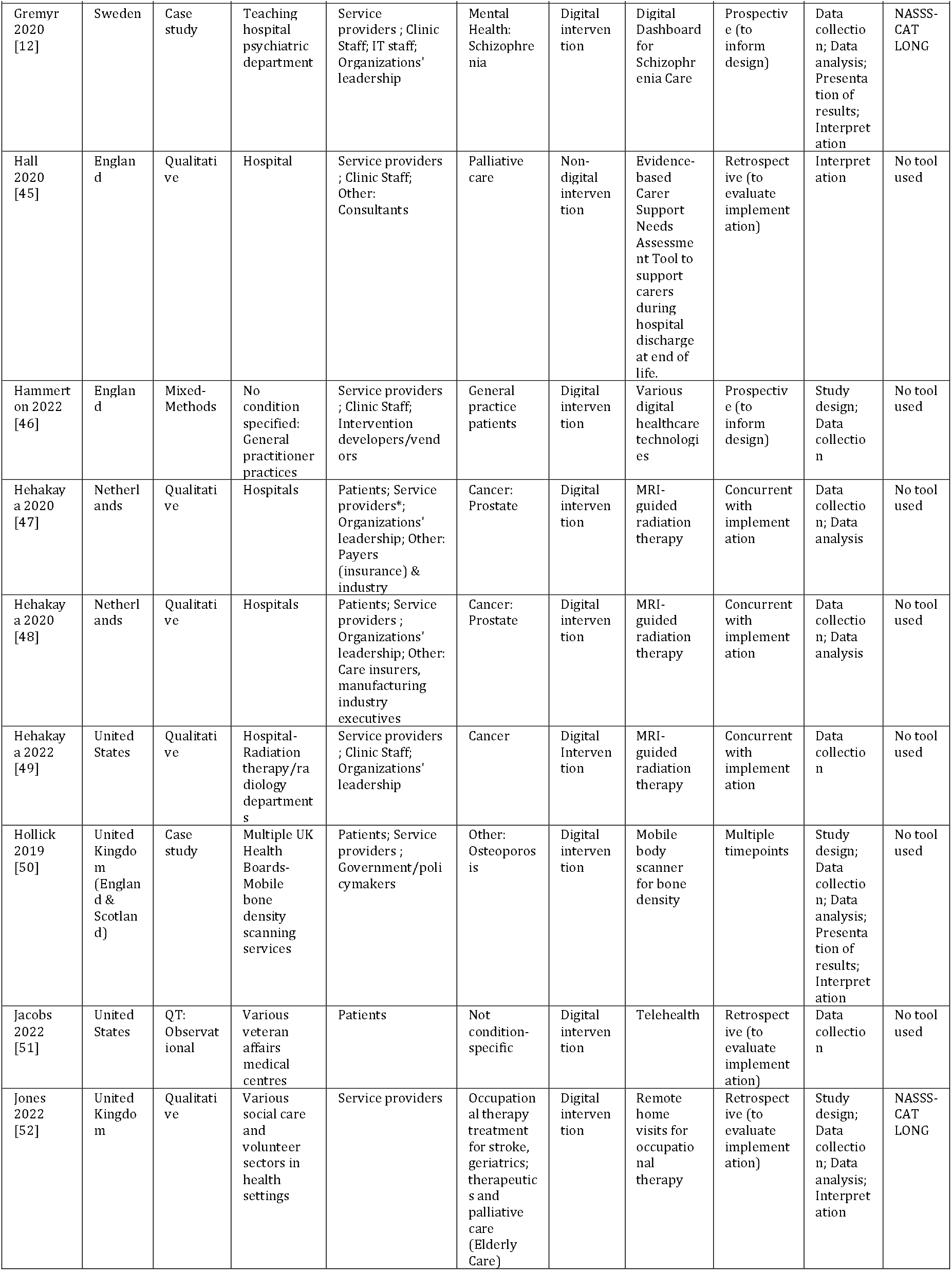
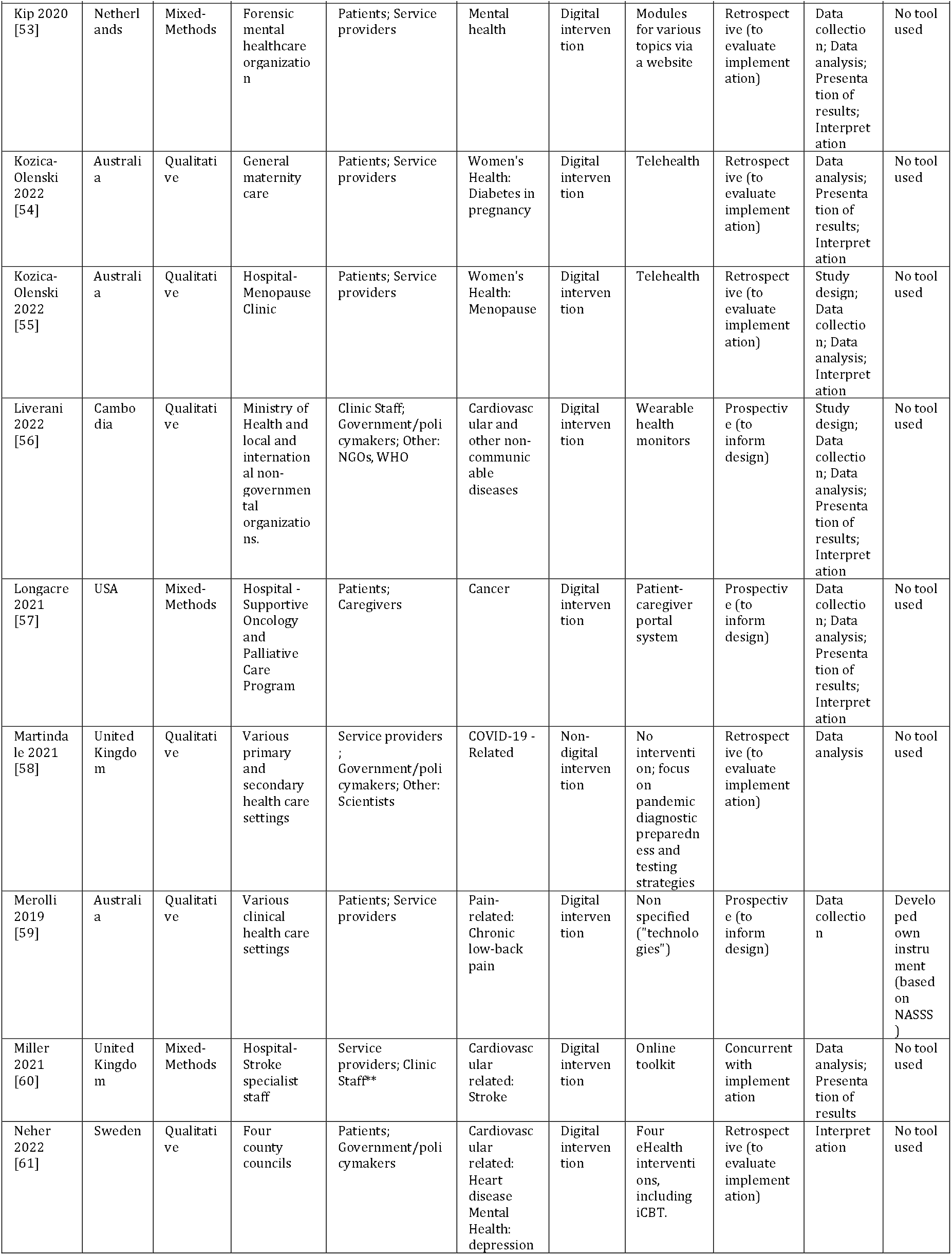
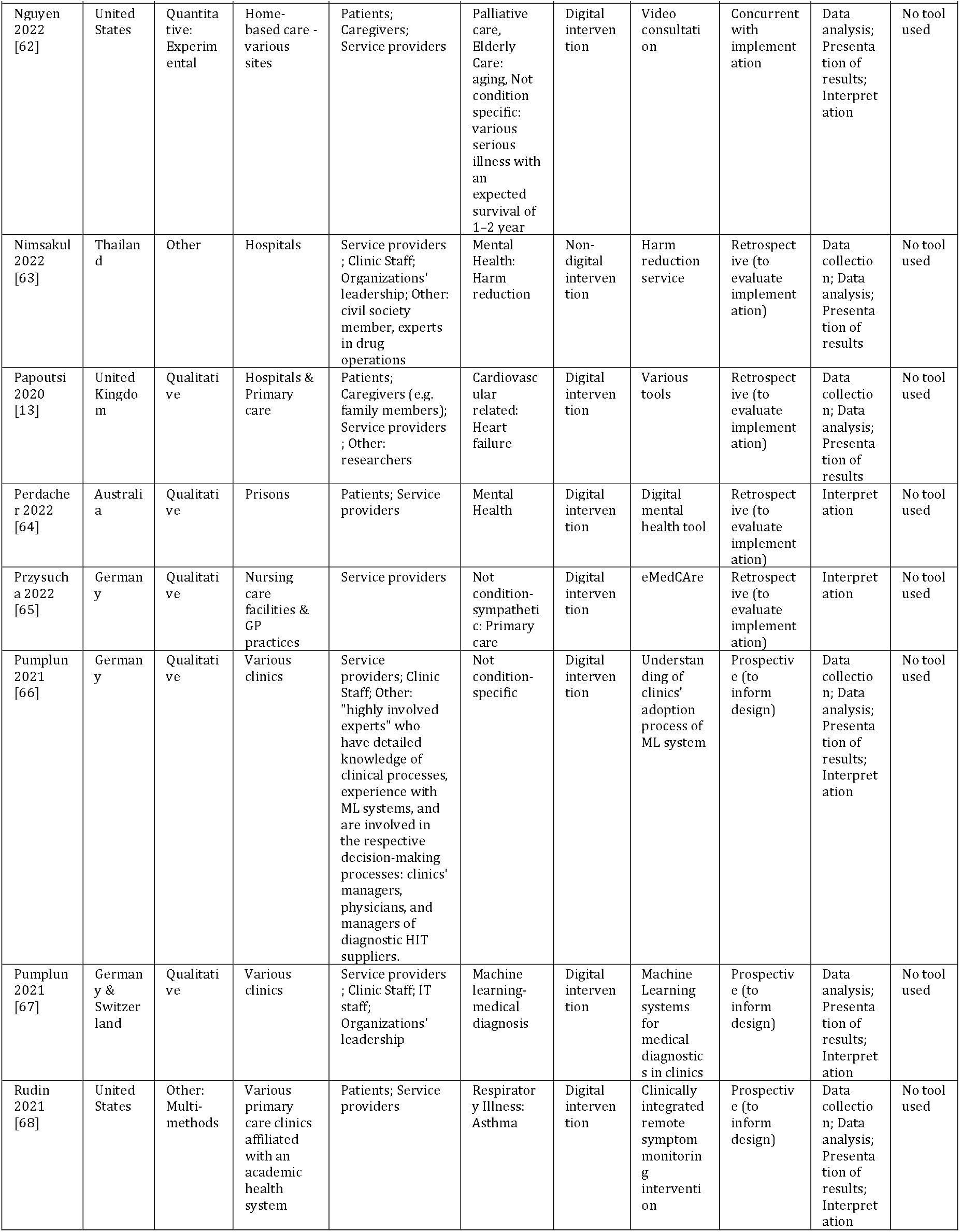
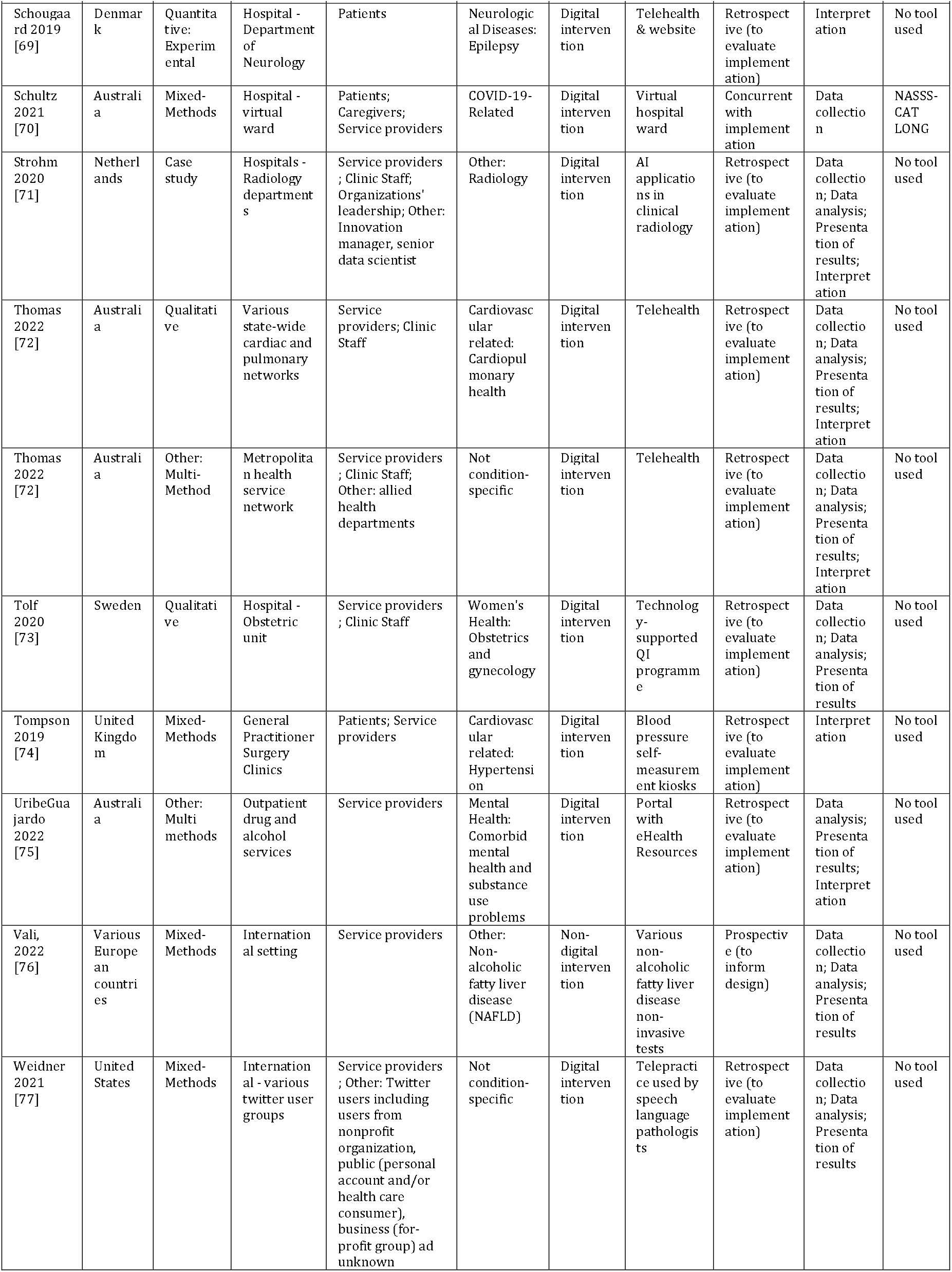
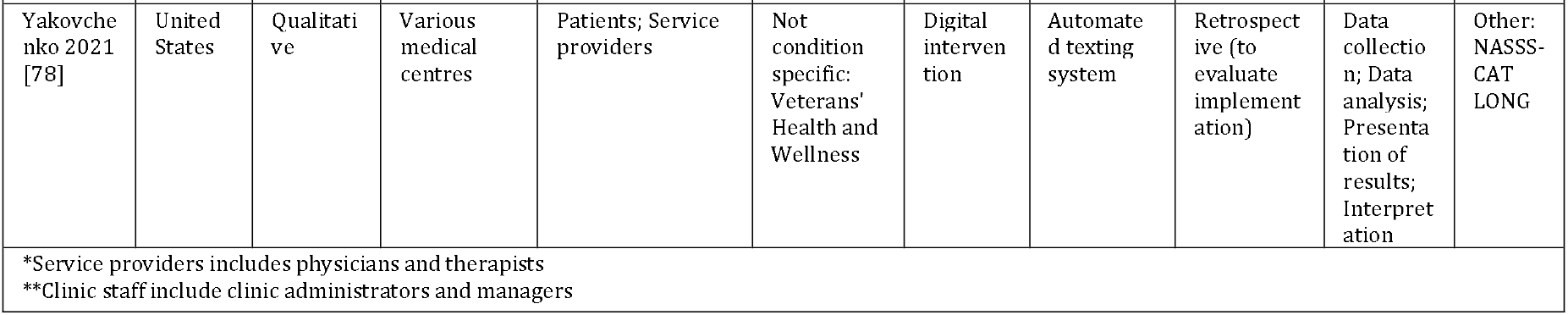
Study characteristics.

**Table 2.** Summary characteristics.

| Characteristic |  |  | Citation |
| --- | --- | --- | --- |
| Study Type | n | % |  |
| *Qualitative* | 28 | 49.1 | [13,27–29,31–33,38,41,42,45,47–49,52,54–56,58,59,61,64–67,72,73,78] |
| *Mixed-methods* | 13 | 22.8 | [26,35,36,40,44,46,53,57,60,70,74,76,77] |
| *Case Study* | 6 | 10.5 | [12,25,34,37,50,71] |
| *Observational* | 3 | 5.3 | [30,43,51] |
| *Experimental* | 3 | 5.3 | [39,62,69] |
| *Other* | 4 | 7.0 | [63,68,75,79] |
| Country of Origin | n | % |  |
| *United Kingdom (including studies set in individual UK countries and across the UK)* | 15 | 26.3 | [13,25,27,31,35,36,39,44–46,50,52,58,60,74] |
| *Australia* | 13 | 22.8 | [26,29,33,40,43,54,55,59,64,70,72,75,79] |
| *USA* | 9 | 15.8 | [41,42,49,51,57,62,68,77,78] |
| *Sweden* | 5 | 8.8 | [12,28,30,61,73] |
| *the Netherlands* | 5 | 8.8 | [37,47,48,53,71] |
| *Germany* | 2 | 3.5 | [65,66] |
| *Thailand* | 1 | 1.8 | [63] |
| *Brazil* | 1 | 1.8 | [34] |
| *Denmark* | 1 | 1.8 | [69] |
| *Canada* | 1 | 1.8 | [32] |
| *Cambodia* | 1 | 1.8 | [56] |
| *Multiple countries* | 3 | 5.3 | [38,67,76] |
| **Conditions Studied\*** | **n** |  |  |
| *Cardiovascular-related* | 10 |  | [13,25–27,33,56,60,61,72,74] |
| *Mental Health* | 9 |  | [12,31,32,39,53,61,63,64,75] |
| *Generalized* | 9 |  | [25,34,41,46,62,65–67,78] |
| *No condition specified* | 5 |  | [36,51,67,77,79] |
| *Cancer* | 5 |  | [44,47–49,57] |
| *Women's health* | 5 |  | [40,44,54,55,73] |
| *Elderly care* | 4 |  | [30,38,52,62] |
| *Neurological disease* | 3 |  | [25,30,69] |
| *COVID-related* | 3 |  | [42,58,70] |
| *Pain-related* | 2 |  | [35,59] |
| *Diet and Nutrition* | 2 |  | [29,43] |
| *Respiratory Illness* | 2 |  | [38,68] |
| *Palliative care* | 2 |  | [45,62] |
| *Other\*\** | 6 |  | [28,37,44,50,71,76] |
| *Not mutually exclusive |  |  |  |
| **Other conditions include diabetes, insomnia, non-alcoholic fatty liver disease, osteoporosis, radiology |  |  |  |

With the NASSS framework having been designed for health technology innovations, there were a variety of health conditions for which innovations were implemented, including cardiovascular (n=10), mental health (n=9), general health promotion (n=9), cancer (n=5), and women’s health (n=5), among others. Of the 57 included studies, 53 implemented digital applications, and the rest (n=4) implemented non-digital interventions such as harm reduction services and COVID-19 testing strategies. Of the 53 digital applications, approximately half of them were telemedicine/virtual care (n=24), followed by personal health devices (n=10), knowledge generation applications (n=9), and digital interventions (n=10), such as internet-based Cognitive Behavioural Therapy (iCBT). See Table 3 for a complete list of digital applications and examples.

**Table 3:**
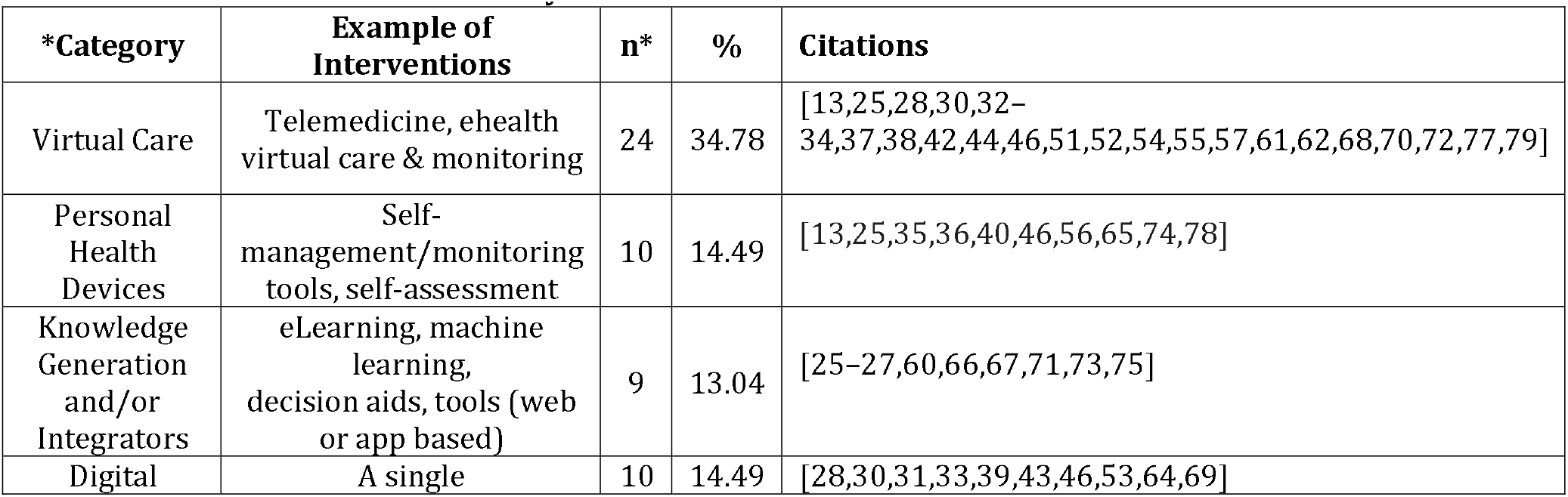
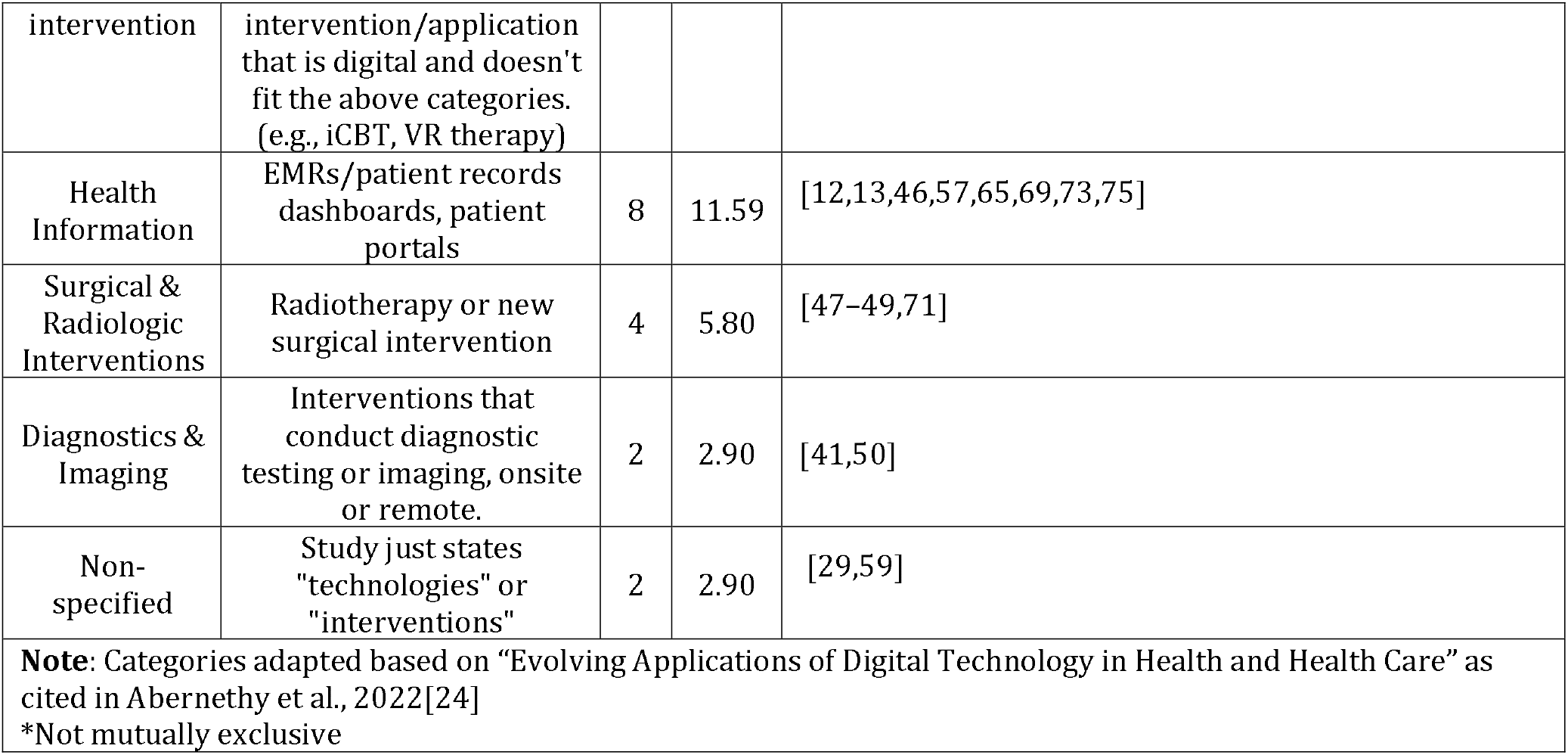
Innovations examined by included studies.

### RQ2. Application of the NASSS framework

As indicated in Table 4, the NASSS framework was used in various aspects of methodology in included studies. The NASSS was used to inform overall study design (n=9), including conceptualization. Studies used the NASSS to inform data collection methods (n=35) by adapting interview guides according to NASSS domains (e.g., [47,72]). Studies also used the NASSS to inform data analysis (n=41), for example by using the NASSS framework for directed content analysis (e.g., [66]). The NASSS was also used to inform data presentation (n=33); studies often utilized a table to organize barriers and enablers by NASSS domain (e.g., [54]). Finally, studies also used the NASSS for interpretation of results (n=39), for example by dedicating one paragraph of the discussion to each NASSS domain (e.g., [61]). Most papers (n=43) used the NASSS to inform multiple aspects of their study.

**Table 4:** Application of the NASSS framework.

| NASSS Application Characteristic | n | % | Citation |
| --- | --- | --- | --- |
| **Study Design Aspect\*** |  |  |  |
| *Overall Study Design* | 9 | 5.7 | [31,37,42,44,46,50,52,55,56] |
| *Data Collection* | 35 | 22.3 | [12,13,27–31,37,38,41–43,46–53,55–57,59,63,67,68,70–73,76–79] |
| *Data Analysis* | 42 | 26.1 | [12,13,25–29,32–40,42,43,47,48,50,52–58,60,62,63,66–68,71–73,75–79] |
| *Presentation of results* | 34 | 21.0 | [12,13,25–29,33–38,42,43,50,53,54,56,57,60,62,63,66–68,71–73,75– |
|  |  |  | 79] |
| *Interpretation of results* | 39 | 24.8 | [12,25–29,31,33–39,42–45,50,52–57,61,62,64–69,71,72,74,75,78,79] |
| **Timing of Implementation** |  |  |  |
| *Retrospectively* | 33 | 57.9 | [13,25–28,32–34,36,37,39–41,45,51–55,58,61,63–65,69,71–75,77–79] |
| *Prospectively* | 15 | 26.3 | [12,29,31,35,38,42,43,46,56,57,59,66–68,76] |
| *Concurrent with Implementation* | 8 | 14.0 | [30,44,47–49,60,62,70] |
| *Multiple Time Points* | 1 | 1.8 | [50] |
| **Number of NASSS domains reported** |  |  |  |
| *1 domain* | 1 | 1.75 | [59] |
| *2 domains* | 1 | 1.75 | [77] |
| *3 domains* | 3 | 5.26 | [43,69,70] |
| *4 domains* | 11 | 19.30 | [27,30,31,35,39,42,46,60,65,74,75] |
| *5 domains* | 9 | 15.79 | [32,34,41,45,48,51,58,61,64] |
| *6 domains* | 13 | 22.81 | [29,44,47,49,52,53,55,57,66–68,71,78] |
| *7 domains* | 19 | 33.33 | [12,13,25,26,28,33,36–38,40,50,54,56,62,63,72,73,76,79] |
| *Not mutually exclusive |  |  |  |

In terms of timing, most studies conducted their analyses using the NASSS framework retrospective to implementation, for example to analyze why implementation did or did not succeed in terms of adoption, non-abandonment, scale, spread, and sustainability of the innovation in a given context (n=33). The rest applied the framework prospectively to inform future implementations (n=15), or concurrently with implementation (n=8). Approximately one third (32%) of included studies reported implementation barriers and enablers related to all 7 NASSS domains, and 21% reported barriers and enablers related to 6 domains. The Embedding and Adaptation Over Time domain was often omitted, but studies incorporated this concept into other domains (e.g., whether the technology will require future iterations [27], whether the regulatory context is expected to change [41]). Another one third (35%) of studies reported barriers and enablers related to four to five NASSS domains, and 12% reported three or fewer, with the latter relying on advisory committees to identify domains of particular relevance to the study [43].

### RQ3. Lessons learned from the application of the NASSS

The barriers and enablers of the successful implementation of innovations are presented by the NASSS domain in Figure 2. The most common barriers across studies (n=47) were in the Organization domain, whereby organizations were cited as lacking in infrastructure, resources, or capacity to innovate and/or whereby the innovation substantially disrupts organizational routines. Specifically, the organization’s capacity, such as technical or human resources, was the most frequently reported barrier and enabler. Another common barrier within the Organization domain was the extent of change required in routines. The following are some exemplary quotes of organizational barriers reported in studies:

> *“Technical infrastructure was sometimes poor, increasing the likelihood of technical crashes”* [26]
>
> *“Representatives from all three groups expressed that an impediment to engaging in the [Quality Improvement] teams was insufficient time and that meeting times conflicted with clinical engagements”* [73]
>
> *“Space and the need for dedicated and private telehealth rooms were also common concerns for clinicians. Such spaces need to be fitted with appropriate hardware, software, and peripheral devices.”* [72]
>
> *“Therapists stated that the intervention was often not discussed in meetings and was not integrated in electronic patient records they used.”* [53]
>
> *“Participants indicated they were concerned that administrative tasks would continue to be a significant time barrier with increased adoption and scale up.”* [29]

**Figure 2:**
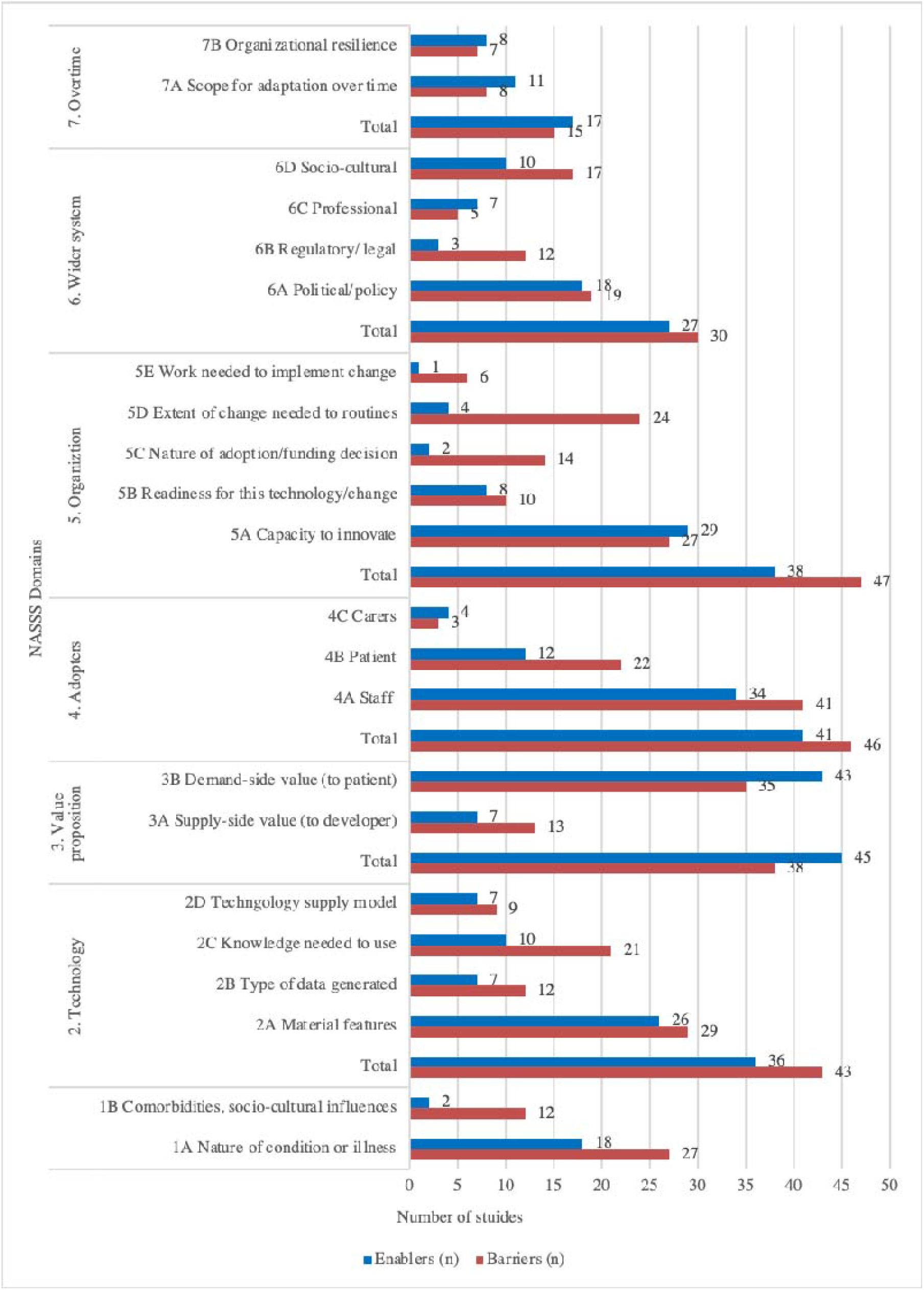
Barriers and Enablers identified in included studies, organized according to NASSS domains.

The most reported enablers were within the Value Proposition domain, whereby a total of 45 studies noted the technology as profitable (from the supply side) or cost-effective (from the demand side) and reported perceived advantages, including improved patient outcomes, increases in access to care, improvements in organizational processes or workflows, and overall effectiveness of the innovation. The following are exemplar quotes of enablers related to the Value Proposition domain reported in studies:

> *“With automated monitoring in the specialist hospital, the accuracy of recording and timely data transfer is reliable. Nurses are more aware of the need to accomplish this task when it’s automated”* [27]
>
> *“Clinicians valued telehealth for the benefits they felt it afforded patients such as convenience and improved access to care, more so than perceived advantages for themselves.”* [55]
>
> *“Several practical advantages were mentioned, among which saving time for therapists and patients because of less traveling time and replacing part of in-person treatment with the intervention, an increase of patients’ access to care because they can individually work on their treatment at their own pace, and providing a new way of delivering treatment to patients.”* [53]

Factors within the Adopter System domain were also commonly reported as barriers or enablers to implementation. A total of 46 studies reported Adopter System factors as barriers, and 41 studies reported them as enablers, including staff, patients, and carers’ attitudes and acceptance towards the new technology and its ease of use. Notably, staff was more frequently reported than patients as both a barrier and an enabler. The following are some exemplar quotes of barriers and enablers related to the Adopter System domain reported in studies:

> *“A few therapists were willing to try ICBT-i, but none were initially deeply interested in the new method, only a few were available to take on this extra task, and only a few had the appropriate competence.”* [28]
>
> *“Lastly, providers described feelings of ‘Zoom fatigue’ and burnout and mentioned that video visits required more concentration, energy, and adaptations to interpret visual cues in comparison to in-person visits”* [32]
>
> *“Most patient participants were interested to see their readings and described the technology as well-designed. They used the tablet and the peripheral devices without too much difficulty and saw great value in monitoring their condition, especially in terms of gaining reassurance and legitimising help-seeking when they needed clinical care.”* [13]

Few authors reported lessons learned from applying the NASSS in their studies. Twenty-five studies commented in varying detail about their experience using NASSS. Eighteen studies [12,26,28,33,35,39,41,50,52,55–57,62,63,68,73,75,78] mentioned that NASSS was a helpful and useful tool, explicitly noting its utility in exploring complexity, facilitating an understanding of the implementation context, applicability in the health technology domain, and its flexibility to be adapted to researchers’ needs. A few studies mentioned the comprehensiveness of the tool for identifying implementation determinants and its value in providing a theoretical foundation [12,27,38]. Additionally, two studies [50,78] suggested future directions for NASSS, such as the opportunity to use the NASSS-CAT tool over time and its applicability in a broader healthcare context. Lastly, two studies [62,67] commented on the limitation of NASSS, including its lack of consideration for how research design can impact intervention implementation and the need for its expansion to include medical ethics.

## Discussion

This scoping review identified 57 empirical studies that used the NASSS framework between its publication in August 2017 and the commencement of the search in December 2022. Most of the included studies were qualitative or mixed/multi-methods designs, which can be attributed to the purpose of NASSS in exploring determinants of implementation success. This exploration required substantial contextual information, and qualitative data could effectively provide it. The NASSS framework was commonly used to inform data collection, data analysis, and the presentation of results. Almost all included studies focused on technological innovation, such as telemedicine/virtual care, health monitoring or decision support via devices and applications, and targeted digital interventions. These innovations were designed for various health conditions (primarily cardiovascular and mental health) or supported general health promotion activities. While approximately one-third of studies reported barriers and enablers for implementation on all 7 NASSS domains, 20% did not report barriers or enablers related to the Over Time domain. The most reported barriers were found in the Organization and Adopter System domains, and the most frequently reported enablers were within the Value Proposition domain.

Most identified studies in this review had used the NASSS retrospectively, primarily to evaluate why an innovation was unsuccessful at becoming adopted by its intended users, got abandoned shortly thereafter, or failed at scaling to become routine within the organization, spreading to other contexts or sustaining over time. Similar findings have been reported with the i-PARiHS (Integrated Promoting Action on Research Implementation in Health Services) application in research [80]. There is a need for prospective and concurrent applications of implementation TMFs to identify potential hurdles and areas of complexity ahead of time with implementation such that mitigation strategies can be put in place [81,82]. Given the novelty of the NASSS framework, many innovations in this review have already been implemented either as small-scale demonstration projects or larger implementations that were not informed a priori by any theoretical framework and therefore required retrospective evaluation. Nevertheless, the NASSS does not offer solutions to identified areas of complexity. While some authors noted that the NASSS helped illuminate areas of focus, it remained unclear what actions they intended to take [26]. A recent companion document, [11], explicitly recommends the next steps for each domain where complexity is identified; however, only four of the included studies had used any of the NASSS-CAT tools [12,52,70,78].

The prevalent implementation determinants (i.e., barriers and enablers) identified in the Organization and Adopter System domains found in this review are consistent with findings in previous reviews of other tools used in implementation science. The Exploration, Preparation, Implementation, Sustainment (EPIS) [83] is a commonly used framework that highlights key phases guiding implementation as well as factors related to the outer (system) context, inner (organizational) context and the innovation itself. A review of this framework application shows that the Implementation phase was most commonly examined in research. During this phase, organizational and individual adopter characteristics were the most frequently mentioned factors [84], as observed in the current NASSS review.

In the dynamic field of implementation science, various determinant frameworks share similarities in understanding complex factors, focusing on contextual elements that influence the successful implementation of healthcare innovations. CFIR, a popular determinant framework in implementation science, primarily identifies factors that influence implementation outcomes across the domains of Intervention Characteristics, Outer Setting, Inner Setting, Individual Characteristics, and Implementation Process [85]. CFIR serves a similar purpose as the NASSS. A recent literature review of CFIR use indicates that the most commonly used constructs in studies were “Knowledge and Beliefs about the Intervention,” followed by “Self-Efficacy,” both of which fall within the domain of Individual Characteristics [86]. This finding aligns with the NASSS’ Adopter System domain and echoes the Value Proposition domain, all commonly reported barriers and enablers in this review.

The i-PARiHS is another implementation determinant framework, and it has four interacting core constructs, including Evidence, Context, Recipients, and Facilitation [87,88]. The inner and outer Contexts in the i-PARiHS are like the Organization and Wider Context domains of the NASSS. A review of research using the i-PARiHS [89] identified variations in how researchers conceptualized outer Context, including specific influences from external organizations, such as guideline-producing entities, and attributions of ’contextual trust’ to broader political and economic characteristics [89]. This conceptualization resonates with the Wider Context of the NASSS. Furthermore, leadership was suggested as another key sub-construct within the Context of the i-PARiHS [89], which corresponds to the 5A Capacity subdomain within the Organization domain of the NASSS.

Although the NASSS was initially created to implement health and care technologies, it exhibits similarities with widely used implementation determinant frameworks designed for a broader range of health innovations, encompassing health technology and evidence-based practices. As such, we found four studies included in this review that used the NASSS for non-digital innovations [45,58,63,76], demonstrating the framework’s adaptability and utility.

Our review found that, when used, the NASSS informed many aspects of design, including the data collection process, data analysis, and the presentation and interpretation of results. The use of the NASSS framework in data collection and analysis was usually consistently and clearly reported. However, there was a lack of consistency and clarity in using the NASSS framework to present and interpret results. Often, data were presented within the primary domains of the NASSS framework. As our team organized narrative descriptions of barriers and enablers into the NASSS subdomains, we observed several instances of overlapping domains.

Furthermore, we identified the potential for these barriers to be mapped onto other primary NASSS domains. This observation may indicate the intricate nature of the implementation under examination in the included studies, which could be explained by the framework’s underlying assumption that, in complex situations, the NASSS domains interact with one another and are interdependent [25]. In other words, when interdependencies among the domains exist, it often leads to the inability to address a singular issue without inadvertently giving rise to new challenges in other domains of the NASSS [25].

For studies that did not present their results using the NASSS domains, despite reporting the NASSS use for data analysis, it became challenging to determine which domain(s) the results pertained to concerning predicting or explaining implementation success or failure. This unclear use of the NASSS framework for presenting results and interpreting findings represents a notable gap in the literature. It has been previously documented in the literature that implementation studies lack reporting, leading to low-quality reporting in the field [90,91].

Specifically, many implementation studies have faced criticism for providing inaccurate descriptions of the context and lacking information detail on the implementation process [91]. Poor reporting makes it difficult to synthesize evidence from relevant studies [90]. Therefore, enhancing reporting practices to facilitate more straightforward evidence synthesis is essential, aiding future empirical testing and refinement of the NASSS.

Additionally, some studies were unclear about how the NASSS framework was used to inform the study designs, including the presentation and/or interpretation of results. Clear reporting standards may increase the NASSS’ utility by guiding researchers on correctly applying and describing its use. The need for better reporting on how TMFs are used in implementation research is a gap in the literature that has already been discussed [92]. For example, in a review of implementation TMFs, 159 different TMFs were identified, with 87% used in five or fewer studies [92]. Despite the substantial number of TMFs, there is limited evidence base describing their use [92]. This limitation restricts opportunities for advancing the science and learning from other researchers. Implementation studies should more clearly report how TMFs have been incorporated into the study design [93]. Better reporting allows for a coherent synthesis of evidence, application and scaling of the TMFs to other contexts, thereby contributing to the science of implementation [93]. We also found that not many authors shared their experience of using the NASSS or provided suggestions for the NASSS advancement. Two studies in this review mentioned the NASSS’ shortcomings [62,67], including ethical principles, and this has been addressed in the Planning and Evaluating Remote Consultation Services framework in 2021[94]. It would be beneficial to conduct a review in five years to reassess the application of the NASSS, explore grey literature, and gather lessons learned for the ongoing advancement and refinement of the framework.

Reporting issues have led to the creation of reporting checklists in other fields, like the Consolidated Standards of Reporting Trials (CONSORT) checklist for randomized controlled trials [95]. Some implementation reporting standards are available; one example is the Standards for Reporting Implementation Studies (StaRI) Statement and Checklist [91]. The StaRI checklist prompts authors to describe the implementation method and the intervention [91], encouraging detailed reporting on contextual information. In addition, the StaRI checklist also prompts authors to describe the theoretical underpinnings of the study. Therefore, its use in future implementation studies is encouraged and may improve reporting of TMF applications, including the NASSS.

### Limitations

Several limitations of this review must be acknowledged. First, quality appraisal was not employed to exclude studies, as scoping reviews generally do not require such assessment. In addition, our primary goal was to explore the breadth and depth of the literature and map available literature about the NASSS application. Second, the field of mHealth is rapidly evolving, and our findings may need re-evaluation.

Nevertheless, our review remains relevant at the time of publication and contributes to the ongoing evolution of the NASSS. Third, this review excluded non-empirical papers, such as commentaries and opinion articles, which could offer authors insights regarding their experiences with the NASSS framework. Future reviews aiming to reassess the NASSS application can include a grey literature search to enhance comprehensiveness. Fourth, we only included studies written in English. While we did include a small number of English studies published in non-English speaking countries, our findings may not provide a comprehensive representation of the NASSS application in those regions.

## Conclusions

This review outlines the characteristics of studies using the NASSS framework and examines patterns of its application. Most of the included studies employed qualitative or mixed/multi-methods designs, which align with the NASSS’s purpose of exploring determinants of implementation success. This often requires qualitative exploration to assess context. Additionally, most studies retrospectively applied the NASSS, likely due to the novelty of the framework. However, this highlights the need for prospective and concurrent utilization of the NASSS during the implementation phase, revealing a gap in the current literature.

Furthermore, nearly all included studies identified various domains as both implementation barriers and enablers, aligning with the current literature on the intricate nature of the implementation process. This underscores the importance of thorough preparation for successful implementation outcomes. Lastly, our review findings point to a need for improved reporting of NASSS utilization in research, including how it was applied and a need for more consistency in presenting results and interpreting findings using the NASSS to facilitate evidence synthesis in the future.

## Data Availability

This is a scoping review of published literature.

## Acknowledgements

HDS conceptualized this review, and HDS, EH, EG, MM, LS, and RB designed the review protocol following the JBI methodology. LS and HDS wrote the review protocol and developed the search strategy with a librarian. HDS, EH, EG, MM, LS, and RB participated in screening titles and abstracts and assessing full texts against the inclusion criteria. HDS, EH, EG, MM, LS, and RB participated in data extraction. HDS, EH, EG, and MM designed the data analysis plan. HDS, EH, EG, MM, LS, and RB participated in data analysis. HDS, EH, EG, MM, KET, and MB participated in data interpretation. HDS, EH, EG, and MM developed tables and figures for data presentation. HDS, EH, EG, and MM wrote the first draft of the review report. All authors critically reviewed and provided feedback on the manuscript. HDS worked on manuscript revisions. KET and MB supervised all steps of this review.

## Abbreviations

NASSS: Non-Adoption, Abandonment, Scale-Up, Spread and Sustainability

## Supporting Information 1. Search Strategy

**(Search Ran 20 December 2022)**

### Medline (Ovid)

Database Ovid MEDLINE: Epub Ahead of Print, In-Process & Other Non-Indexed Citations, Ovid MEDLINE® Daily and Ovid MEDLINE® <1946-Present>

Search strategy:

1. NASSS.ti,ab,kf. (51)
2. ((non-adoption or nonadoption) adj2 abandonment adj5 (scale-up or scaleup) adj2 spread adj2 sustainability).ti,ab,kf. (53)
3. NASSS-CAT.ti,ab,kf. (2)
4. (greenhalgh* adj5 (framework* or model*)).ti,ab,kf. (26)
5. 1 or 2 or 3 or 4 (88)

### EMBASE (Ovid)

Database: Embase Classic+Embase 1947 to 2022 December 19 Search strategy:

1. NASSS.ti,ab,kf. (54)
2. ((non-adoption or nonadoption) adj2 abandonment adj5 (scale-up or scaleup) adj2 spread adj2 sustainability).ti,ab,kf. (51)
3. NASSS-CAT.ti,ab,kf. (2)
4. (greenhalgh* adj5 (framework* or model*)).ti,ab,kf. (35)
5. 1 or 2 or 3 or 4 (97)

### APA PsychInfo (Ovid)

Database: APA PsycInfo 1806 to December Week 2 2022 Search strategy:

1. NASSS.ti,ab,id. (12)
2. ((non-adoption or nonadoption) adj2 abandonment adj5 (scale-up or scaleup) adj2 spread adj2 sustainability).ti,ab,id. (6)
3. NASSS-CAT.ti,ab,id. (0)
4. (greenhalgh* adj5 (framework* or model*)).ti,ab,id. (8)
5. 1 or 2 or 3 or 4 (22)

### CINAHL (EBSCO)

Database: CINAHL Plus with Full Text Search strategy:

1. TI NASSS OR AB NASSS OR TX NASSS (34)
2. TI ( ((non-adoption OR nonadoption) N2 abandonment N2 (scale-up or scaleup) N2 spread N2 sustainability) ) OR AB ( ((non-adoption OR nonadoption) N2 abandonment N2 (scale-up or scaleup) N2 spread N2 sustainability) ) OR TX ( ((non-adoption OR nonadoption) N2 abandonment N2 (scale-up or scaleup) N2 spread N2 sustainability) ) (20)
3. TI NASSS-CAT OR AB NASSS-CAT OR TX NASSS-CAT (3)
4. TI ( greenhalgh* N5 (framework* OR model*) ) OR AB ( greenhalgh* N5 (framework* OR model*) ) OR TX ( greenhalgh* N5 (framework* OR model*) ) (74) 5 S1 OR S2 OR S3 OR S4 (109)

### LISTA (EBSCO)

Database: Library, Information Science & Technology Abstracts Search strategy:

1. TI NASSS OR AB NASSS OR TX NASSS (9)
2. TI ( ((non-adoption OR nonadoption) N2 abandonment N2 (scale-up or scaleup) N2 spread N2 sustainability) ) OR AB ( ((non-adoption OR nonadoption) N2 abandonment N2 (scale-up or scaleup) N2 spread N2 sustainability) ) OR TX ( ((non-adoption OR nonadoption) N2 abandonment N2 (scale-up or scaleup) N2 spread N2 sustainability) ) (9)
3. TI NASSS-CAT OR AB NASSS-CAT OR TX NASSS-CAT (1)
4. TI ( greenhalgh* N5 (framework* OR model*) ) OR AB ( greenhalgh* N5 (framework* OR model*) ) OR TX ( greenhalgh* N5 (framework* OR model*) ) (1) 5 S1 OR S2 OR S3 OR S4 (12)

### Web of Science

Database: Web of Science Core Collection (1900-present) Search strategy:

1. TS=(NASSS) (48)
2. TS=((non-adoption or nonadoption) NEAR/2 abandonment NEAR/5 (scale-up or scaleup) NEAR/2 spread NEAR/2 sustainability ) (49)
3. TS=(NASSS-CAT) (2)
4. TS=(greenhalgh* NEAR/5 (framework* OR model*)) (30)
5. #1 OR #2 OR #3 OR #4 (89)

### Scopus

Database: Scopus Search strategy:

1. TITLE-ABS-KEY ( nasss ) (55)
2. TITLE-ABS-KEY ( ( NON-ADOPTION OR NONADOPTION ) W/2 ABANDONMENT W/5 ( SCALE-UP OR SCALEUP ) W/2 SPREAD W/2 SUSTAINABILITY ) (43)
3. TITLE-ABS-KEY ( nasss-cat ) (2)
4. TITLE-ABS-KEY ( greenhalgh* W/5 ( framework* OR model* ) ) (38)
5. TITLE-ABS-KEY ( nasss ) ) OR ( TITLE-ABS-KEY ( ( non adoption OR nonadoption ) W/2 abandonment W/5 ( scale-up OR scaleup ) W/2 spread W/2 sustainability ) ) OR ( TITLE-ABS-KEY ( nasss-cat ) ) OR ( TITLE-ABS-KEY ( greenhalgh* W/5 ( framework* OR model* ) ) ) (103)

## Supporting Information 2. Data Extraction Tool

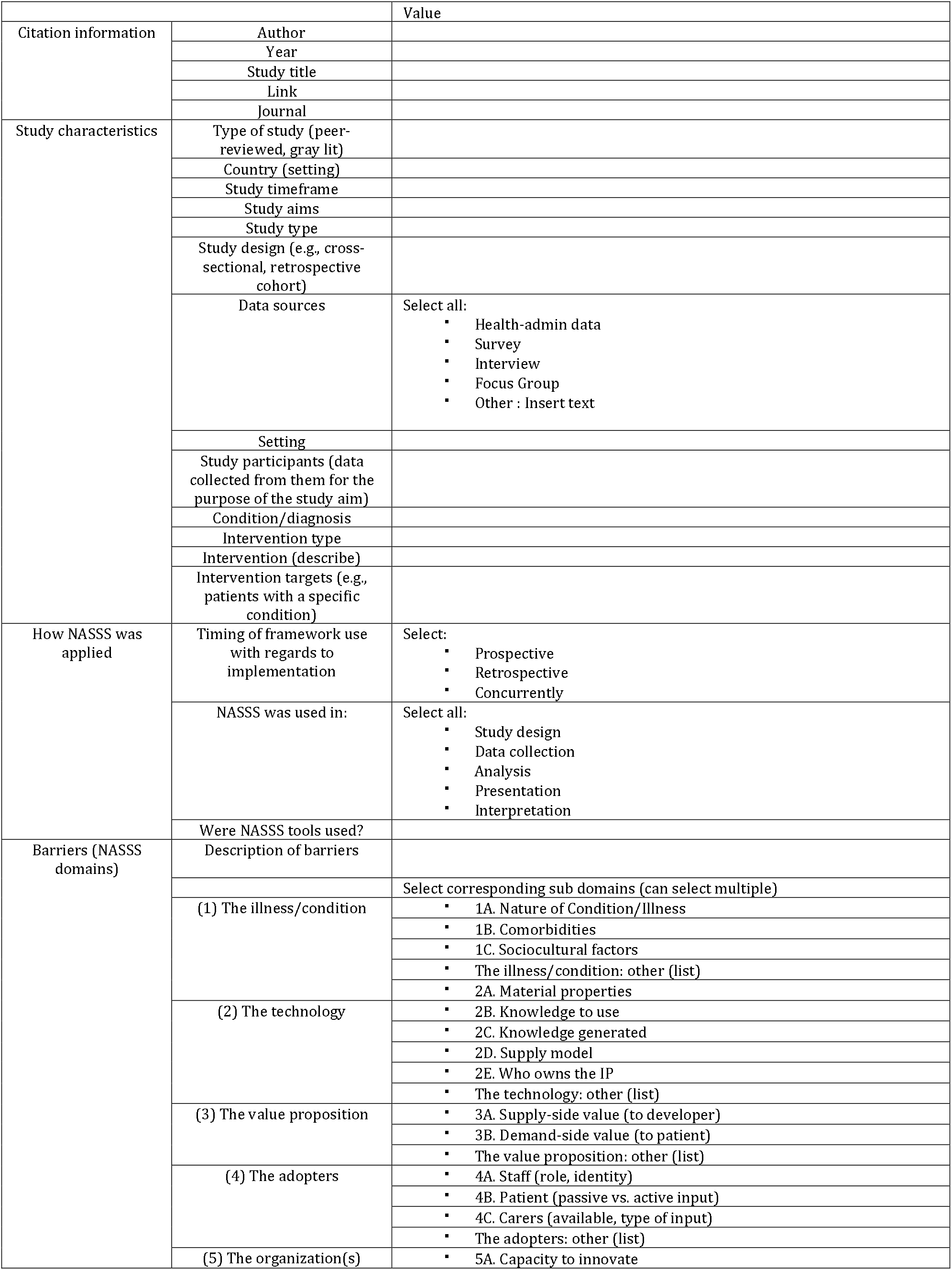
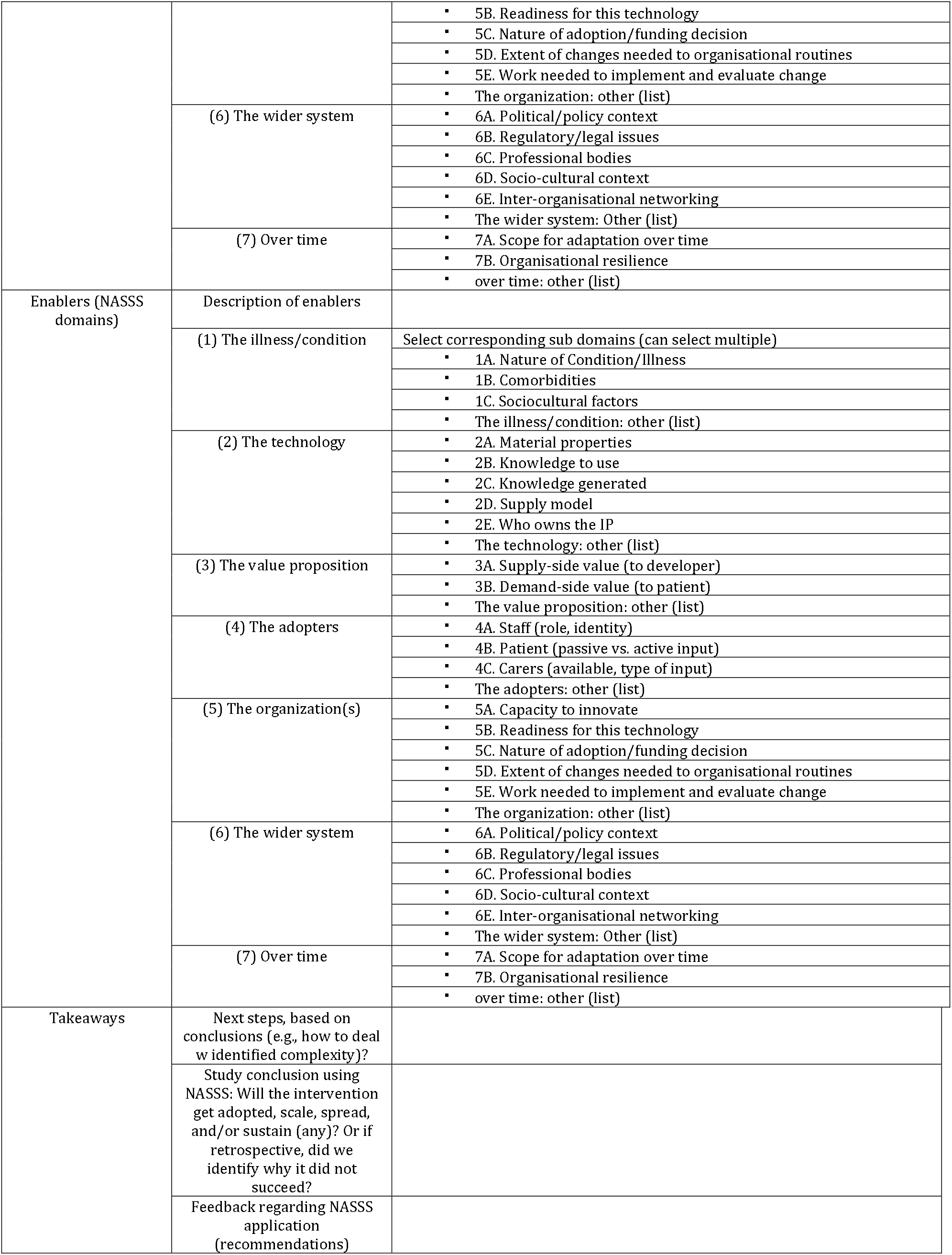

